# Projecting Demand-Supply Gap of Hospital Capacity in India in the face of COVID-19 pandemic using Age-Structured deterministic SEIR model

**DOI:** 10.1101/2020.05.14.20100537

**Authors:** Veenapani Rajeev Verma, Anuraag Saini, Sumirtha Gandhi, Umakant Dash, Muhammad Shaffi Fazaludeen Koya

## Abstract

**BACKGROUND:** Due to uncertainties encompassing the transmission dynamics of COVID-19, mathematical models informing the trajectory of disease are being proposed throughout the world. Current pandemic is also characterized by surge in hospitalizations which has overwhelmed even the most resilient health systems. Therefore, it is imperative to assess supply side preparedness in tandem with demand projections for comprehensive outlook.

**OBJECTIVE:** Hence, we attempted this study to forecast the demand of hospital resources for one year period and correspondingly assessed capacity and tipping points of Indian health system to absorb surges in demand due to COVID-19.

**METHODS:** We employed age-structured deterministic SEIR model and modified it to allow for testing and isolation capacity to forecast the demand under varying scenarios. Projections for documented cases were made for varying degree of mitigation strategies of a) No-lockdown b) Moderate-lockdown c) Full-lockdown. Correspondingly, data on a) General beds b) ICU beds and c) Ventilators was collated from various government records. Further, we computed the daily turnover of each of these resources which was then adjusted for proportion of cases requiring mild, severe and critical care to arrive at maximum number of COVID-19 cases manageable by health care system of India.

**FINDINGS:** Our results revealed pervasive deficits in the capacity of public health system to absorb surge in demand during peak of epidemic. Also, continuing strict lockdown measures was found to be ineffective in suppressing total infections significantly, rather would only push the peak by a month. However, augmented testing of 500,000 tests per day during peak (mid-July) under moderate lockdown scenario would lead to more reported cases (5,500,000–6,000,000), leading to surge in demand for hospital resources. A minimum allocation of 10% public resources and 30% private resources would be required to commensurate with demand under that scenario. However, if the testing capacity is limited to 200,000 tests per day under same scenario, documented cases would plummet by half.

## 1. INTRODUCTION

Coronavirus disease 2019(COVID-19) is a contagious disease caused by a novel strain of coronavirus (SARS-CoV-2) which was first informed as the cluster of viral pneumonia cases of unknown etiology detected in Wuhan City, Hubei Province, China. Coronaviruses are enveloped, positive single-stranded large RNA virus that infect humans, but also a wide range of animals (1). Amongst humans, they have known to cause myriad of illness with varying degree of severity ranging from relatively benign common cold to more severe forms like SARS (Severe Acute Respiratory Syndrome) and MERS (Middle East Respiratory Syndrome). However, illness onset among rapidly increasing number of people and mounting evidence human-to-human transmission suggests that SARS-CoV-2 is more contagious than its predecessors (2).

Following the outbreak of COVID-19, the WHO Emergency Committee declared it a global health emergency on January 30, 2020 and a pandemic on March 11, 2020. Within a short span of time, a localized outbreak evolved into pandemic with three defining characteristics: a) Speed and Scale-the disease has spread quickly to all corners of the world, and its capacity for explosive spread has overwhelmed even the most resilient health systems b) Severity-Overall, 20% cases are severe or critical, with a crude clinical case fatality rate currently of over 3%, increasing in older age groups and in those with certain underlying conditions c) Societal and economic disruption-shocks to health and social care systems and measures taken to control transmission having deep socio-economic consequences (3). Currently, approximately 4,217,408 confirmed cases of COVID-19 has been reported including an estimated 284,736 deaths in 210 countries and territories as on May 11, 2020.

Around three months have elapsed since the first case of COVID-19 was reported in India on January 30, 2020. Since then, the number of cases have surged to 69,149 with 2,249 deaths as on May11, 2020. As part of the pandemic preparedness, in the absence of vaccines or antivirals, India adapted gamut of non-pharmaceutical interventions such as lockdown, quarantining and tracing and testing concomitantly to alter the trajectory of pandemic. These interventions encompassed two types of strategies a) Suppression, which aims to reverse epidemic growth, by minimizing the effective reproduction number (average number of secondary cases each case generates), R, to below 1 and thus, reduce case numbers to low levels and maintaining that indefinitely b) Mitigation, which aims to slow the spread of epidemic with the rationale of preventing significant overload on health system and gradually allowing the population to develop herd immunity (4).

WHO enforced a binding instrument of International Health Regulation (IHR) in 2007 to prevent, detect and respond to public health emergencies. The IHR monitoring and evaluation framework includes State Party Annual Reporting Tool (SPAR) which underscores 13 capacity indicators to gauge the preparedness of nations to mitigate the effect of public health emergencies, including the emergence of novel pathogen (WHO, 2018). In 2018, although India’s SPAR composite score (0.75) was above the international average (0.61), albeit, the scores for indicators pertaining to health service provisioning and laboratory capacity were incongruous as India had only half the average readiness in these indicators as compared to international scores. Therefore, it is imperative to unravel the supply side readiness especially, with regards to infrastructural capacity of India to handle the surge of hospitalization cases.

Demand for hospitalization services for COVID-19 can be estimated by analyzing the interface between transmission curve, age-structure, contact patterns and morbidity status of population. Disease progression is characterized by myriad of uncertainties and the trajectory of an epidemic is defined by some key factors and parameters. Specifically, for a novel infection whose disease dynamics are still unclear, mathematical models are thus, pertinent to understand the mechanics of transmission. Deterministic compartmental models such as Susceptible-Exposed-Infectious and Recovered (SEIR) are widely used to provide insight into disease progression and can be chosen over complex models due to minimum number of assumptions. Yet, there is a caveat in using the baseline SEIR model as it doesn’t incorporate the testing capacity in the model which punctuates the dynamics in two ways. Firstly, in basic model, the undetected yet infectious individuals are not accounted in determining probability of infection and potential transmissibility and secondly, there’s reduced transmissibility from confirmed positive cases which no longer transmit the disease once they are tested and isolated. Modelling exercises allowing for testing and isolation capacity and undetected cases are rather scarce. Further, COVID-19 has differential impact on different age groups and additionally, heterogeneities in contact networks have a major effect in determining whether pathogen can become epidemic or persist at endemic levels. Consequently, mathematical models of disease transmission incorporating age and social contact structures are more congruous to the reality.

Therefore, in this study we have attempted the short and long term prediction of transmission of COVID-19 using age-structured compartment based model allowing heterogeneities in contact networks and simulating for varying assumptions and scenarios around containment and mitigation strategies. Drawing from international experience and literature on COVID-19, we determined the proportion of high-risk population with underlying conditions in India who are more vulnerable to progress into severe condition upon infection that can guide triage and targeted intervention decisions. Further, we assessed the capacity and tipping points of Indian health system to absorb surges in the number of people that will need hospitalization and critical care because of COVID-19 based on varying scenarios.

## 2. OBJECTIVES

The objectives of the study can be elucidated as follows: – a) Estimate the projected demand for hospital resources under various mitigation strategies of reduced social mixing and varying levels of testing capacity in India. b) Analyze the hospital surge capacity of the Indian Health System to absorb the surge in demand under different scenarios.

## 3. METHODS

### 3.1 Demand Estimation

#### 3.1.1 Model

The spread of any virus is incumbent upon the infectivity of pathogen and the pool of susceptible population. We formulated the transmission dynamics model for the outbreak of COVID-19 in a heterogeneously mixing population. Compartmental disease models divide population into groups (or compartments) based on each individual’s infection status and track the corresponding population sizes through time. SEIR model in which the population was divided according to infection status into Susceptible(S), Exposed (E), Infected (I) and Removed(R) was employed in the study. Susceptible individuals become infected at a given rate when they interact/contact with an infectious person and enter the exposed state. These exposed pool transition to infectious state after a latency period and later either recover or die. However, it is crucial to consider host age structure to enable realistic modelling for disease prevention policy. Also, the spread of an infectious disease is sensitive to the contact patterns in the population as person-to-person transmission is largely driven by who interacts with whom. The age-specific mixing patterns of individuals in age group *i* alter their likelihood of being exposed to the virus given the extent of infections in the group. The assumption of homogenously mixing population can lead to an overestimation of the final epidemic size and magnitude of interventions needed to stop an epidemic(5). Contrarily, including contact patterns that vary across age and locations (e.g. home, work, and schools) as predictors in transmission dynamic model improves the model’s realism. Thus, we investigated the impact that different mixing assumptions have on the spread of COVID-19 in an age structured SEIR model with infectivity in both latent and infectious period described by a system of ordinary differential equations.

#### 3.1.2 Model Map

Our model incorporates two sections as illustrated in Figure 1 where left part represents the classic SEIR model with additional post latency node accounting for infections in the terminal stage of incubation period. The transmission dynamics is further branched out into asymptomatic, symptomatic, severe and critical on the right-hand side constrained by the testing coverage. Following equations represents the dynamics of the model.

**Figure 1.**
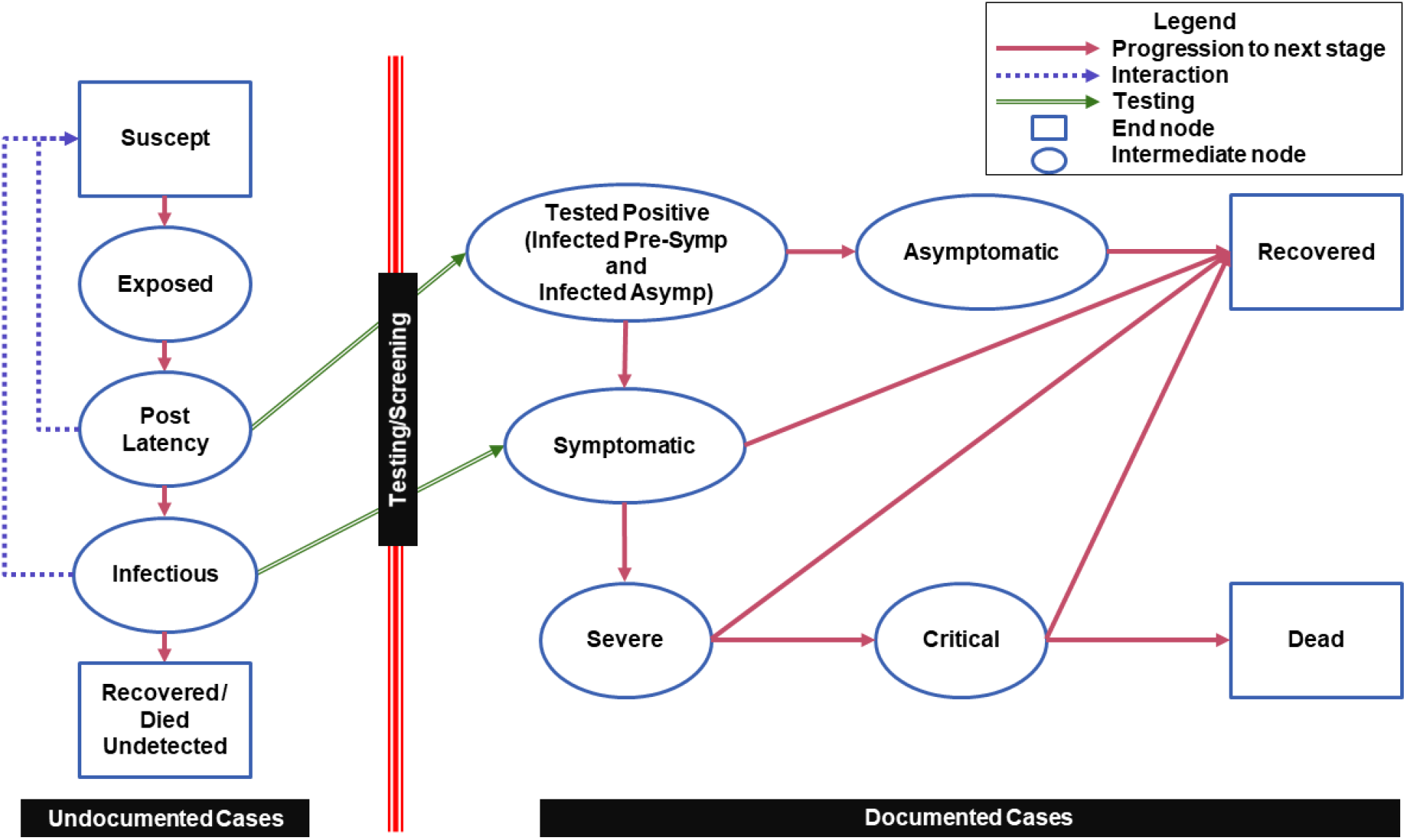
Model Map

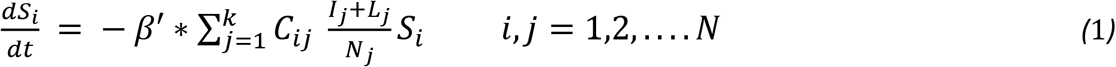

Where β’ is the probability of infection upon contact with infectious people, N is the total number of considered age groups, C_ij_ is the number of age wise interaction between infectious and susceptible population.

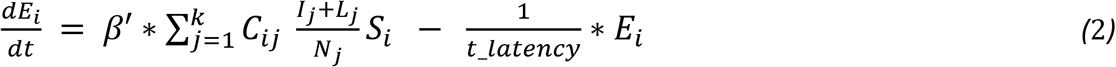

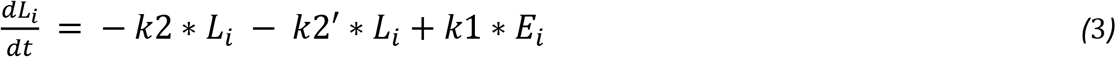

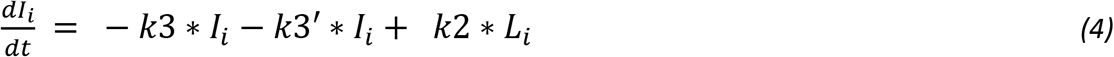

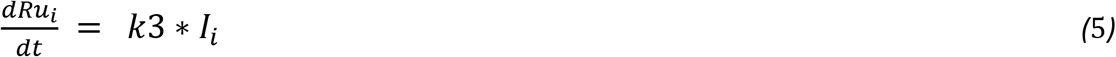

Where S_i_, E_i_, L_i_, I_i_ and Ru_i_ are the susceptible, exposed, post latency, infectious and undocumented recovered group of people of age group *i*. Rate constant k1 = 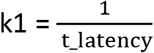 is defined by time from exposure to onset of infectiousness (latent period), k2’ and k3’ are rate for testing, k2 is defined by part of incubation time after latent period and k3 is rate at which undocumented infectious people recover or die.

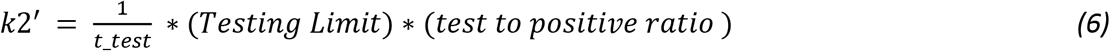

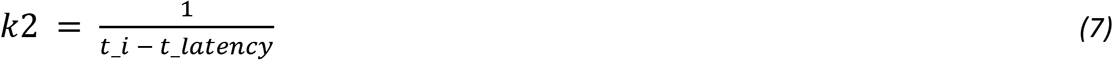

Documented cases in the model are determined and constrained by the testing. Positive cases can be Presymptomatic, Symptomatic and Asymptomatic. Pre-symptomatic and Symptomatic may progress to severe and critical state before either recovery or death. Whereas true asymptomatic positives recover without exhibiting any symptoms over the course of illness.

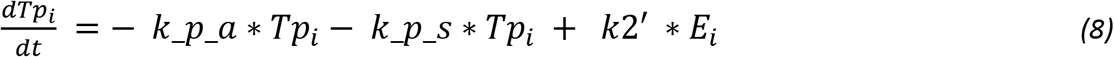

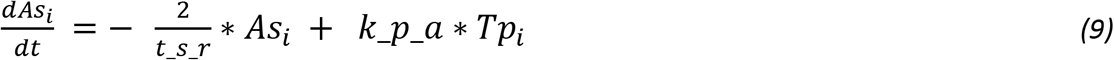

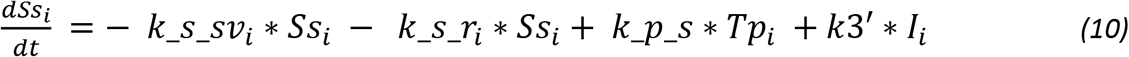

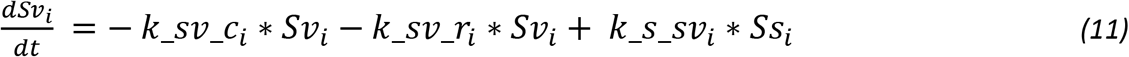

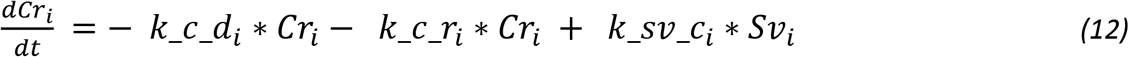

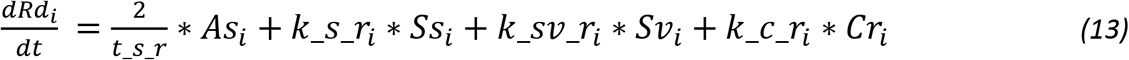

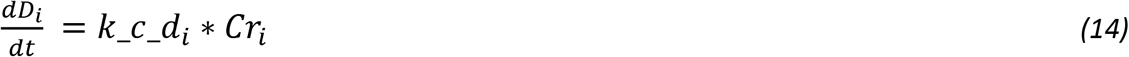

Where Tp_i_ is tested positive, As_i_ is asymptomatic, Ss_i_ is symptomatic, Sv_i_ is severe, Cr_i_ is critical, D_i_ is dead and Rd_i_ is documented recovered for age group i. Rates of progression are given as follows

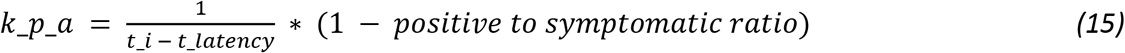

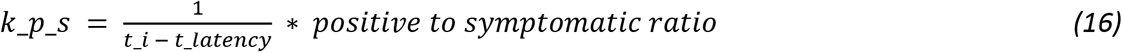

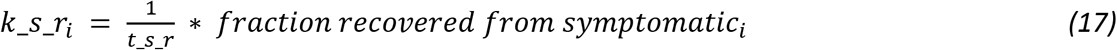

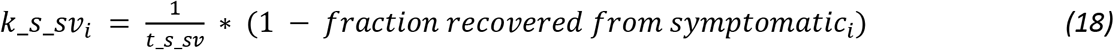

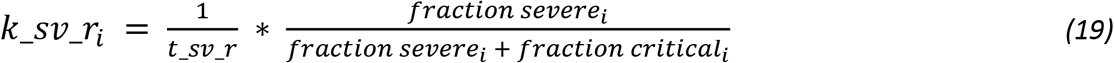

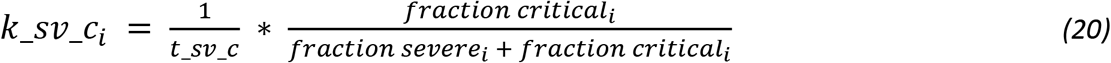

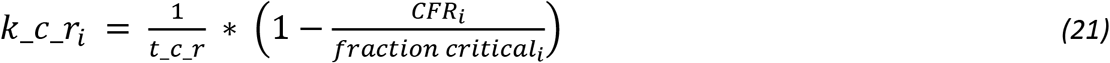

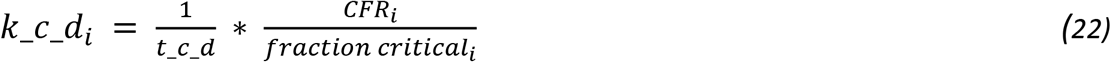

To account for the mitigation factor such as social distancing, the contact matrix is divided into household contacts, workplace contacts, school contacts and other contacts using the following form

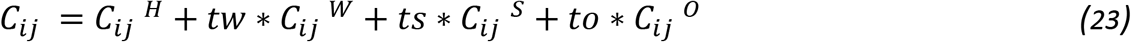

Where *tw, ts* and *to* can range from 0 to 1 allowing various degree of interactions of contacts delineating the impact of same on progression of the epidemic.

#### 3.1.3 Model parameters and data

Main parameters of the model are surmised in Table 1 which were extracted/estimated from various sources. Population age distribution for India was obtained from website of Population Pyramid (6) and Social Contact Matrices were adapted from state-of-the-art compilation by Prem et. al in which matrices were projected by Bayesian hierarchical model for 152 countries using contact surveys and demographic data (7).Time parameters were extracted from a recently published systematic review which synthesized parameters using meta-analysis of 43 studies on COVID-19 (8). Further, age-distributed data from U.S. was used to estimate the epidemiological parameters in the study. As reported in Table 2, age wise fractions of transition from one state to another and case fatality ratio (CFR) was calculated from the report published by U.S. Center for Disease Control (9), as currently in India (as of 2^nd^ May), CFR is analogous to that of USA when CDC carried out the study. Also, we collated the data on confirmed positive cases, hospitalizations, recovery and deaths from publicly available time series data of USA published by U.S. CDC(10) to estimate the probability of infection β’. Remaining model parameters such as latency rate, testing rate and rate of recovery/death were estimated using crowdsourced Indian data (11) from 10^th^ March 2020 till 2^nd^ May, 2020 while incorporating mitigation strategies of lockdown starting from 24^th^ March to 3^rd^ May and current social distancing guidelines based on the zones which is issued by Indian government starting from 3^rd^ May to 17^th^ May. Three scenarios of mitigation measures were modeled – (a) Full lockdown (assuming closure of schools, workplace and community spaces and doubling of contacts in households) (b) Social distancing measures and moderate lockdown (assuming closure of schools, staggered opening of workplace and community spaces with half strength and social distancing of vulnerable population including aged above 70 years and people with at-least one underlying high risk chronic condition with 75% compliance) post 17^th^ May, 2020 and (c) No lockdown (assuming 100% contacts in schools, workplace, community spaces and households). The model is augmented to incorporate the testing coverage and Test to positive (*TTP*) ratio of 5% till May 2^nd^ in India.

**Table 1.** Parameters used in the model equations

| Name | Parameter | Value | Reference |
| --- | --- | --- | --- |
| Incubation time | $t_i$ | $5.83 \pm 0.515$ | Meta-analysis (8) |
| Symptomatic to Recover time | $t_{s_r}$ | $11.76 \pm 2.61$ | |
| Symptomatic to Severe time | $t_{s_{sv}}$ | $4.82 \pm 0.683$ | |
| Severe to Recover time | $t_{sv_r}$ | $17.76 \pm 2.61$ | |
| Sever to Critical time | $t_{sv_c}$ | $5.66 \pm 0.765$ | |
| Critical to Recover time | $t_{c_r}$ | $17.76 \pm 2.61$ | |
| Critical to Dead time | $t_{c_r}$ | $5.45 \pm 1.49$ | |
| Testing time | $t_{test}$ | 1 | Estimated from crowdsourced Indian data (https://www.covid19india.org/) |
| Latency time | $t_{latency}$ | 3 | |
| Rate of recovery/death | $k_3$ | 0.1 | |
| Test to positive ratio | - | 5% |  |
| Probability of infection | $\beta'$ | 0.11 | Estimated from U.S data (https://www.cdc.gov) |
| Positive to pre-symptomatic/symptomatic ratio | - | 98% | ECDC (24) |

**Table 2.**
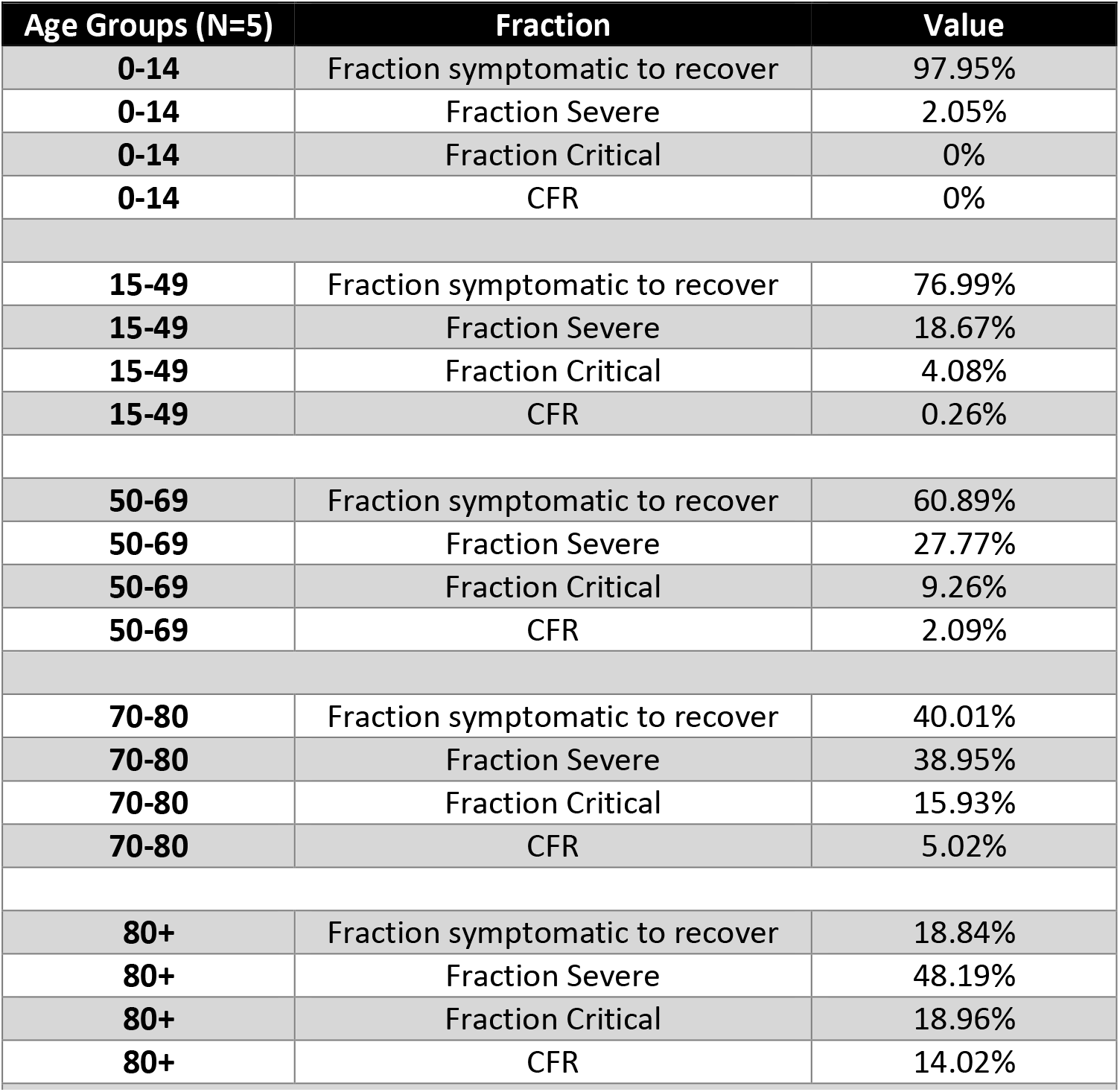
Age wise fractions in symptomatic, severe, critical, dead and recovered

| Age Groups (N=5) | Fraction | Value |
| --- | --- | --- |
| 0-14 | Fraction symptomatic to recover | 97.95% |
| 0-14 | Fraction Severe | 2.05% |
| 0-14 | Fraction Critical | 0% |
| 0-14 | CFR | 0% |
| 15-49 | Fraction symptomatic to recover | 76.99% |
| 15-49 | Fraction Severe | 18.67% |
| 15-49 | Fraction Critical | 4.08% |
| 15-49 | CFR | 0.26% |
| 50-69 | Fraction symptomatic to recover | 60.89% |
| 50-69 | Fraction Severe | 27.77% |
| 50-69 | Fraction Critical | 9.26% |
| 50-69 | CFR | 2.09% |
| 70-80 | Fraction symptomatic to recover | 40.01% |
| 70-80 | Fraction Severe | 38.95% |
| 70-80 | Fraction Critical | 15.93% |
| 70-80 | CFR | 5.02% |
| 80+ | Fraction symptomatic to recover | 18.84% |
| 80+ | Fraction Severe | 48.19% |
| 80+ | Fraction Critical | 18.96% |
| 80+ | CFR | 14.02% |

#### 3.1.4 Model Assumptions

The underlying set of assumptions characterizing the transmission dynamics are elucidated as follows-

i. India was assumed to be a closed system with constant population size of 1.3 billion (S+E+L+I+R = 1.3 billion) throughout the course of epidemic.
ii. Different inflows and outflows and imbalance by demographics, migration etc. is not considered.
iii. Seasonal effects, weather conditions like temperature and humidity and mutations not included in the model.
iv. Heterogeneous mixing in the contact networks is assumed and interactions at four spheres are considered: home, work, school and other locations.
v. Infectivity from latency period is considered, a latent individual can transmit the disease to the susceptible, i.e., individual has force of infection in both latent and infectious period.
vi. Asymptomatic transmission from truly asymptomatic cases is accounted in the model.
vii. Positively diagnosed cases are documented and isolated from rest of the susceptible pool, thus, less likely to spread the infection.
viii. The recovered individuals develop the immunity and do not revert back to the susceptible pool.

### 3.2 Proportion of high-risk individuals

Multi country evidence suggest that there are certain underlying conditions most associated with severe and critical cases of COVID-19. In conjunction with elderly, younger and working population group with chronic health conditions are also vulnerable and can be classified as high risk for developing complications. U.S. Center for Disease Control and Prevention in its Morbidity and Mortality Weekly report (12)divulged that as of March 28, 2020, 71% hospitalized non-ICU cases and 78% ICU cases had one or more precondition. Similarly, evidence from Italy (13) affirmed that in a sample of 355 deaths in Italy,99.2% patients had some pre-condition(s). Overall, among dead, 25.1% had single disease, 25.6% had 2 diseases and 48.5% had 3 or more underlying diseases in Italy. Therefore, analyzing the prevalence of high-risk underlying condition associated with COVID-19 is germane to understand the progression of cases from mild to severe/critical stage and identify the vulnerable population to design more effective interventions. Hence, we used the information on burden of chronic diseases from a study conducted by London School of Hygiene and Tropical Medicine (14) in which number of individuals at increased risk of severe COVID-19 disease were estimated for 188 countries. The study identified following underlying conditions characterizing high-risk groups and special populations requiring urgent care- (a) HIV/AIDS (b) Tuberculosis (c) Cancers with direct immunosuppression (d) Cancers with possible immunosuppression (e) Cardiovascular diseases (f) Chronic respiratory diseases (g) Cirrhosis and other chronic liver diseases (h) Diabetes mellitus (i) Chronic kidney diseases (j) Chronic neurological disorders (h) Sickle cell disorders. Therefore, we used the information on prevalence of these underlying conditions in India (Table 3) by culling out information from above-mentioned study to estimate proportion of population with at-least one underlying condition.

**Table 3.**
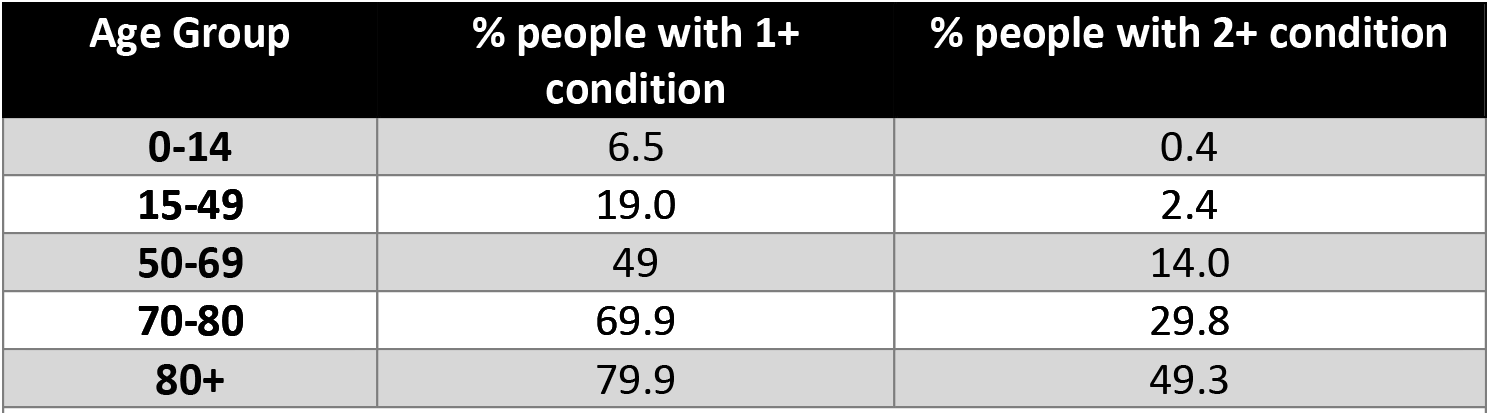
Fraction of India’s population with 1+ and 2+ underlying conditions

| Age Group | % people with 1+ condition | % people with 2+ condition |
| --- | --- | --- |
| 0-14 | 6.5 | 0.4 |
| 15-49 | 19.0 | 2.4 |
| 50-69 | 49 | 14.0 |
| 70-80 | 69.9 | 29.8 |
| 80+ | 79.9 | 49.3 |

### 3.3 Estimating health system surge capacity

Data on Indian health infrastructure was coalesced from different sources. Information on beds in public hospitals (PHC+CHC+DH) and Medical College Hospitals was extracted from National Health Profile, 2019(15). However, there was missing information in this data source for some states and colleges, which was then scrapped individually from websites of the colleges. Beds under AYUSH, Defense, Railways and ESI Corporation are also incorporated in the analysis as India is likely to rope in beds from these institutions during surge in demand. The data for bed availability in these institutions was taken from National Health Profile, 2018(16). Further, due to absence of veritable data available on private sector, approximations using utilization rates for hospitalizations in private hospitals from recently released National Sample Survey Organization 75^TH^ round (17) was used and crude estimations were made based on proportion of inpatients treated in private hospitals. Further, there is major lacunae in credible data availability for ICU beds and Ventilators required for critical care. Henceforth, another assumption was made that only 5% of total beds are ICU beds and 50% ICU beds are equipped with ventilators in India.

Health system’s capacity to accomodate the increasing number of COVID-19 patients was computed using an open access tool developed by Giannakeas et al (2020) (18). This tool enables the modelling of steady state patient flow dynamics that can guide the tipping point of health care system in terms of the availability of hospital beds, ICU beds and mechanical ventilators. Entire dynamics of supply side readiness is explained in Figure 4. The left-hand panel in the figure is representative of the current health capacity with total number of hospital beds, ICU beds and ventilators. Average length of stay determining the daily turnover rates for mild, severe and critical cases were set as 11.76 days, 17.76 days and 17.76 days respectively (parameter values from model). Using population-weighted age-stratified probabilities, number of cases requiring hospital beds, ICU beds and ventilators was estimated. Further, the daily turnover of each of these resources was measured by dividing the number of available resources with average length of stay for that resource. Thereafter, the daily turnover rates were divided by the proportion of cases requiring mild, severe and critical care to arrive at the maximum number of COVID cases manageable by health care system of India. Three distinct cases based on the allocation of resources were made to discern the surge capacity for each of these cases. In all three cases, we assumed that public health facilities dedicate fixed 10% of their hospital beds, ICU beds and ventilators for COVID-19 patients since existing bed occupancy rate of public hospitals in India is around 94% (19). However, for private sector hospitals, varied assumptions pertaining to allocation was made. Under case 1, the provision of private infrastructure was assumed to be 0. Whereas, the allocation of hospital beds, ICU beds and ventilators in private hospitals was 10% under case 2 and further expanded to 30% under case-3

## 4. RESULTS

Main findings of the study are presented in this section. Figure 1 encapsulates various transmission curves under the scenario of ramping up of testing coverage (number of individuals tested daily) and increased *TTP* from 5% to 20% when the pandemic reaches final stage of community transmission. The impact of other non –pharmaceutical interventions like social distancing and lockdown is also explicated in the figure. While the lockdown was effective in slowing down the infection rate and shifting the peak to later months of the year 2020, it perpetuated heavy socio-economic costs in India. Therefore, it is pertinent to compare how continuing with moderate lockdown or lifting the restrictions completely would impact the number and spread of total covid-19 cases in India.

### 4.1 Projected demand for hospital resources

The detailed representation of our modelling results is exhibited in Figure 2. Our modelling exercise revealed that with the current testing rate of 50,000 individuals tested daily on an average in India from 10^th^ March, 2020 till 2^nd^ May 2020 and 5% *TTP* ratio, the estimated total documented covid-19 infections in India would not exhibit any significant difference (300,000 to 600,000 in an year) across the scenarios of no lockdown, full lockdown and moderate lockdown guidelines. Lifting the lockdown completely after 17^th^ May would hasten the surge in demand to 20^th^ July as compared to 23^rd^ August if the full lockdown continues for one year. Although, moderate lockdown would push the peak to early August, total number of infections will not plummet by this intervention.

**Figure 2.**
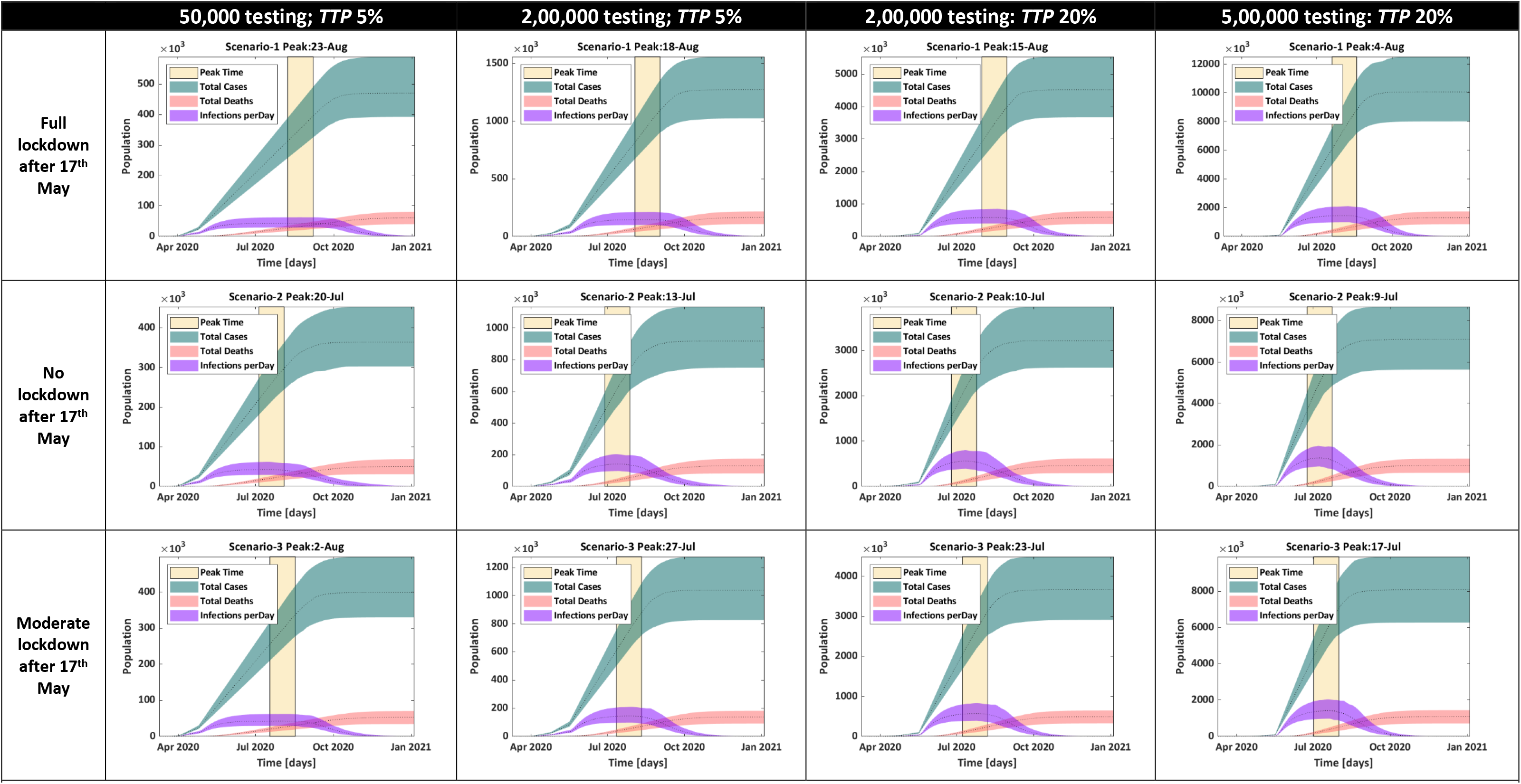
Scenarios of full lockdown, no lockdown and moderate lockdown compared with varying testing rates for India post Lockdown 3.0

Current *TTP* ratio in India is around 5% which is exiguous as compared to *TTP* in USA of 20%. There could be plethora of reasons for low *TTP* in India such as absence of widespread-community spread, untargeted testing or less sensitivity of diagnostic kits. However, we estimated the number of total infections and peak time for surge in demand for expanded testing coverage of 200,000 and 500,000 tests per day at current 5% *TTP* as well as for 20% *TTP* modelling for worst case scenario.

Assuming no/limited community spread with *TTP* of 5% and augmenting the testing to 200,000, in the absence of lockdown, our model estimated the peak to be around mid-July with total documented cases ranging from 500,000 to 700,000 during peak time. However, continuing moderate lockdown (with 50% contacts in schools, workplaces and others), estimated number of documented cases would be around 800,000 at the peak time of 27^th^ July. With this increased testing of 200,000 individuals per day, surge in demand would be highest (∼1,000,000) if stringent lockdown measures are imposed after 17^th^ May. This is because complete lockdown will delay the overall community spread and more people will be diagnosed and tested positive during peak time.

Under the worst case scenario of widespread community spread in India with TTP of 20%, peak will be witnessed around early July 2020 to mid-August 2020. Total documented infections under moderate lockdown scenario will range from 2,500,000 to 3,000,000 around 23^rd^ July if 200,000 individuals are tested daily. However the total documented infections will increase with higher testing rate of 500,000 individual per day ranging from 5,500,000 to 6,000,000 cases around peak time on 17^th^ July. In a no lockdown situation during worst case scenario with TTP 20%, number of cases will peak a week ahead around 10^th^ July with similar number of infected cases in moderate lockdown.

### 4.2 Estimating Surge Capacity of Indian Healthcare System

Our analysis indicated paucity of resources in India to handle surge in demand during the peak time for both mild infections and severe cases. The capacity of Indian healthcare system to absorb increases in caseload is constrained by availability of beds and ventilators. India has a total (public +private) bed capacity of 2,363,296 which can be allocated for COVID-19 infections according to existing bed occupancy rate and government guidelines. The availability of health care infrastructure which is collected from multiple sources is presented in Table 4. According to Case-1 where no allocation is made by private sector and only 10% resources are dedicated in public infrastructure, actual availability of hospital beds, ICU beds and ventilators is 101,131, 5,322 and 2,661 respectively. The estimated inflow/outflow of mild, severe and critical cases per day are 9193, 266 and 133. Under this scenario, India has the capacity to accommodate only 33,870 mild cases, 1,067 severe and critical new COVID cases every day. However, leveraging upon expansive private health care system in India can bolster the capacity. The tipping point for beds and ventilators increases significantly when 10% (case 2) and 30% (case 3) private health care resources were assumed to earmarked to accommodate COVID-19 patients. For example, under case-2, we found that the surge capacity of health system for mild, severe and critical care is distinctly higher than case 1 as can be seen in Figure 3 and Figure 4. Moreover, when the allocation of private infrastructure increases by 30%, the capacity to manage new COVID cases doubles.

**Table 4.**
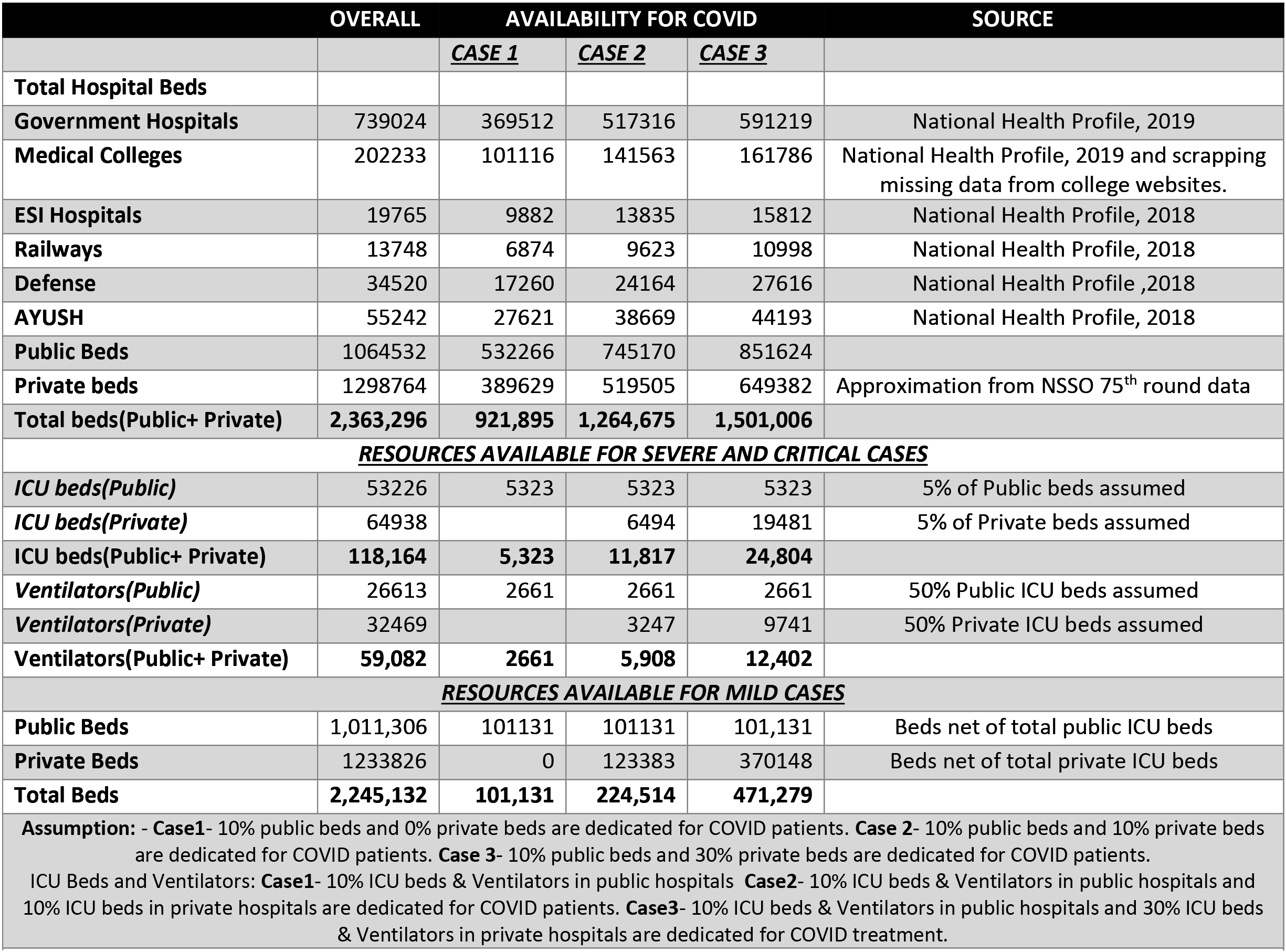
Number of hospital beds, ICUs and ventilators available in India

**Figure 3.**
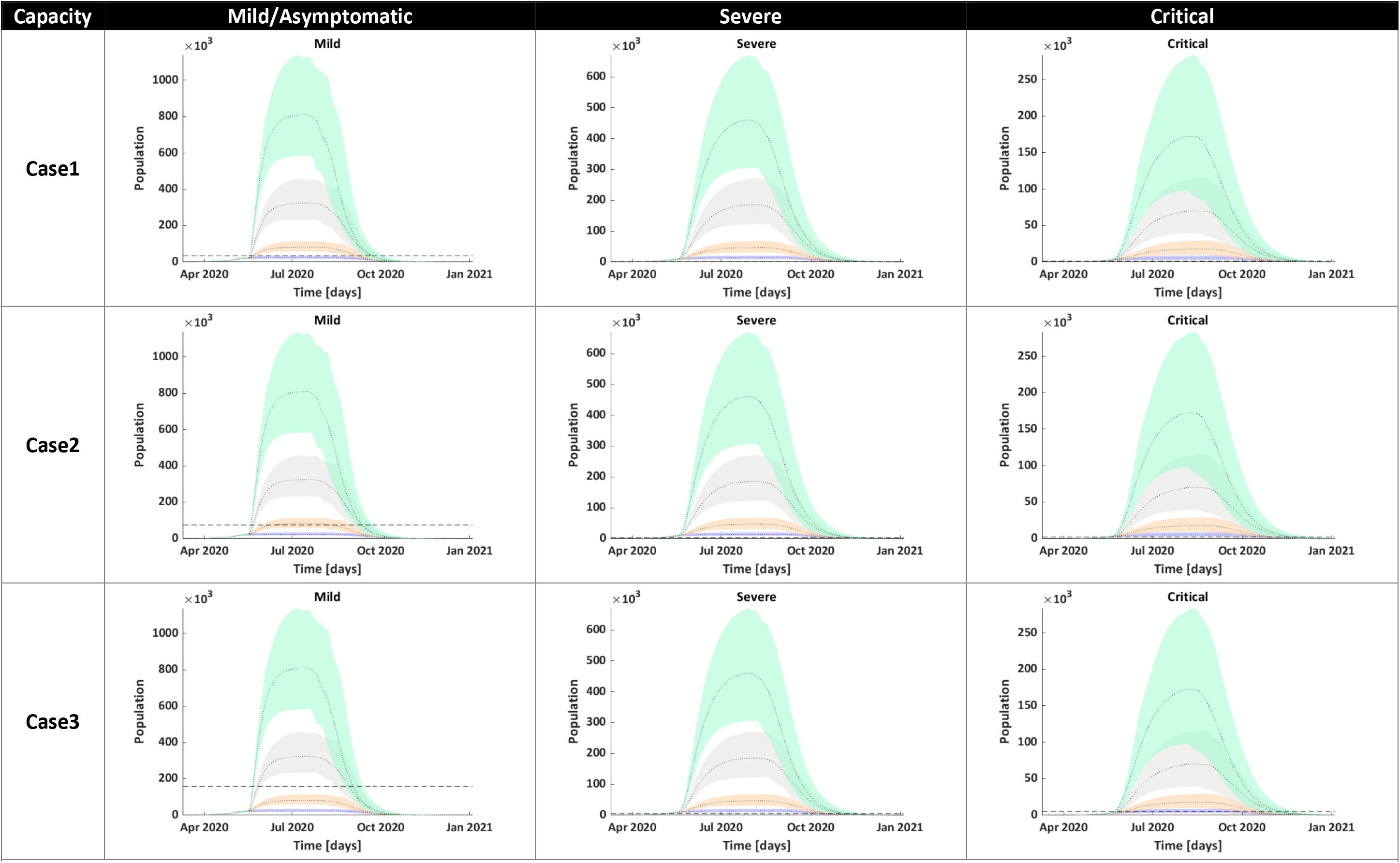
Assesing Demand for varying level of severity under Moderate Lockdown Blue: 50,000 tests with 5% ttp; Peach: 2,00,000 tests with 5% ttp; Grey: 2,00,000 tests with 20% ttp; Cyan: 5,00,000 tests with 20% ttp *ttp denotes test to positive ratio

**Figure 4.**
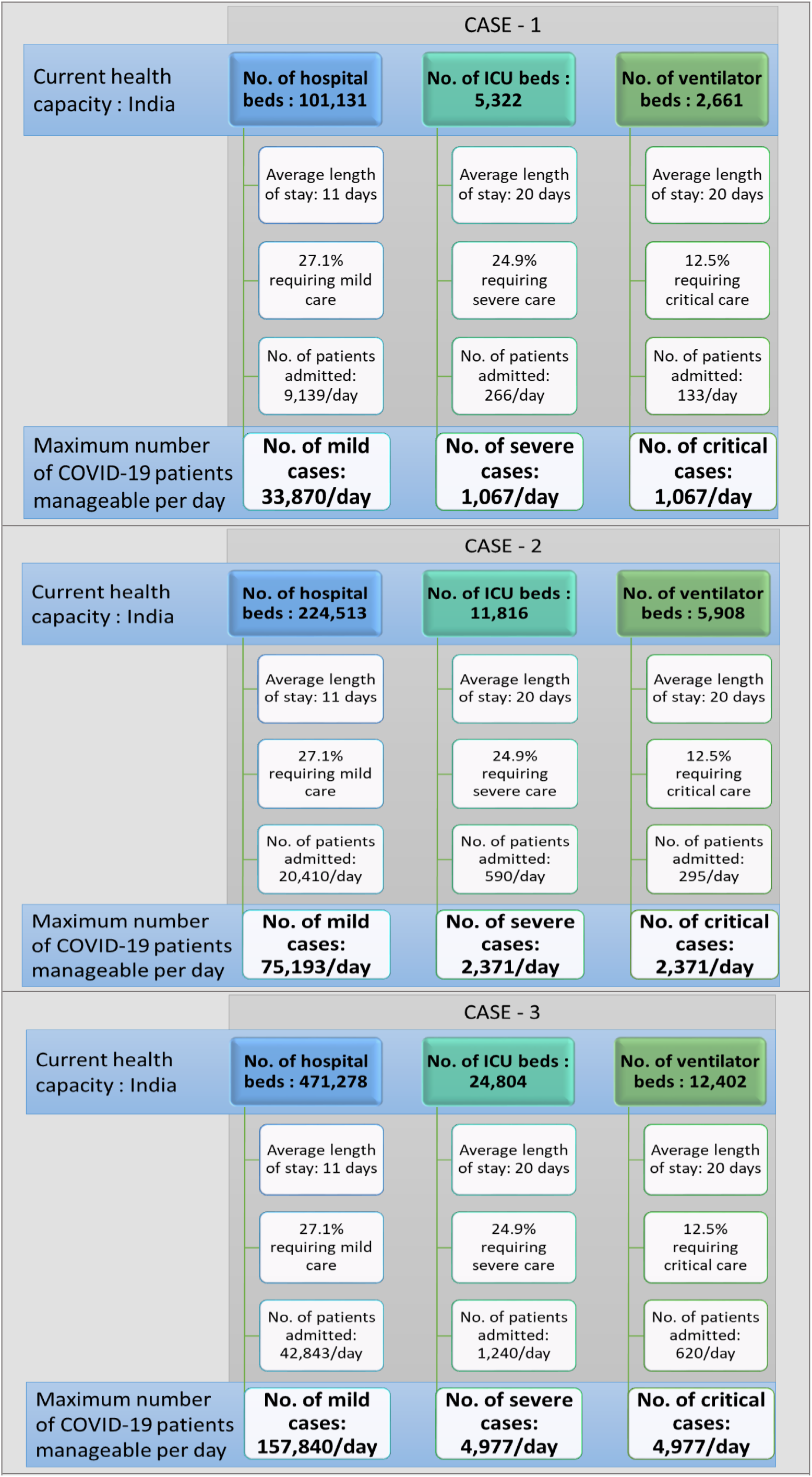
Maximum daily number of incident COVID-19 cases manageable by healthcare system

### 4.3 Demand-Supply Gap under various scenarios

We computed the surge capacity, which is determined by trajectory of cases and infrastructure dedicated to COVID-19 patients by accessing the time period around which Indian hospitals will face serious crisis in terms of availability of beds and ventilators. The demand-supply gap during peak time if the moderate lockdown is continued after 17^th^ May for mild, severe and critical cases is presented in the form of heat map (Figure 5). Green coloured boxes are indicative of the availability of infrastructure during peak time in respective scenarios, whereas non-green coloured boxes suggest the extent of health infrastructure that has to be upgraded to meet the real demand during peak time. As mentioned earlier, surge capacity is likely to vary across different testing scenarios. For example, when the partial lockdown is considered with increased testing coverage of 200,000 and 500,000 per day and *TTP* of 5%, our demand and supply side projections indicate that the Indian health care system is likely to face a huge deficit in terms of the availability of health care infrastructure, especially for severe and critical case.

**Figure 5.**
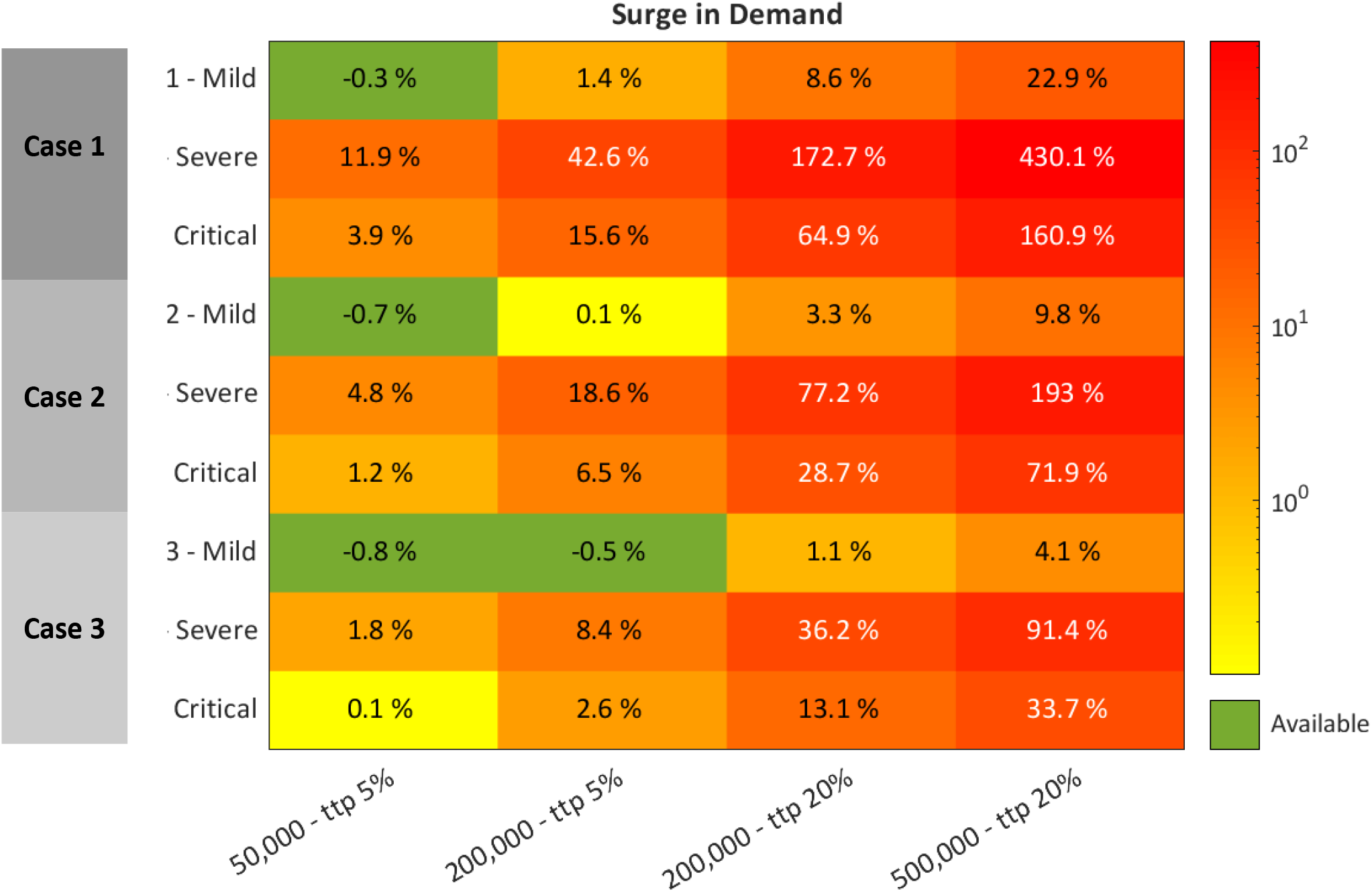
Percent beds available (green) and needed (non-green) from maximum capacity during peak time if moderate lockdown continues with multiple increased testing scenarios. Refer Table 4 for cases definitions

Assuming the community spread and scaling up of testing to 500,000 tests per day with *TTP* of 20%, under Case-1, India will have to increase the availability of ICU beds by more than five times (430%) and availability of ventilators by 2.5 times(150%) before 17^th^ July 2020. However, under current scenario where *TTP* is 5%, if we increase testing to 200,000 cases per day, we will have to ramp up the availability of ICU beds by 42.6% and ventilators by 15.6% before 27^th^ July 2020. Under case-3, where 10% of capacity in public facilities and 30% in private facilities is apportioned for COVID-19, and testing coverage is 200,000 per day with *TTP* of 5%, the estimated demand for severe and critical cases can only be met if supply of ICU beds and ventilators is increased by 8.4% and 2.6% before 27^th^ July 2020. However, increasing the testing even further to 500,000, number of ICU beds must double, and supply of ventilators must be increased by 33.7% before 17^th^ July. Therefore, necessary steps must be made to include private-sector hospitals in the treatment of COVID-19 patients in order to be prepared for the widespread community transmission and increase in detected cases due to aggressive testing. Time to surge in capacity or tipping point will occur much earlier than the peak. Hence, this analysis delineates an urgent need of procuring the ventilators and upgrading the capacity in this 2 months window.

## 5. DISCUSSION

Our study calibrated the model to preliminary data arising from outbreak in India in order to project the demand for hospital resources under three transmission curve scenarios: No lockdown, Moderate lockdown and Full lockdown across varying testing coverage. We also evaluated the extent to which the full lockdown and moderate lockdown delays the peak of outbreak, thereby, prolonging the window of time to augment the health-system capacity in order to accommodate the surge in demand during peak period. Our analysis of Indian healthcare system’s preparedness to absorb surges for infected cases exhibited pervasive deficits. There was a substantial variation in tipping points of supply side capacity across the assumptions on resource allocation in private sector to accommodate COVID-19 patients.

One of the important finding of our analysis is relative ineffectiveness of further extending the strict lockdown measures, as full lockdown is likely to push the surge in demand for hospital resources by a month without suppressing the total number of cases significantly. India was amongst the countries to implement lockdown early with highest stringency in the world which is also indicated by the stringency index of lockdown (100% for India) prepared by University of Oxford (20). Due to the high costs of lockdowns, there’s a policy conundrum if the countries should quarantine everyone at a large social cost or test everyone and apply quarantine in a more directed fashion. The increase in documented cases with augmented testing in our study suggests that lockdown should be replaced gradually with more thrust on public health intervention of testing, tracing and isolating in conjunction with surveillance and real time data. More targeted sequestering of infected case(both symptomatic and asymptomatic) aided with increased random testing, along-with the social distancing measures for containment zones with higher incidence of cases and isolating vulnerable with underlying conditions and aged population is recommended rather than stringent lockdown in order to prevent the negative shock from deepening further.

Our analysis underscored the absence of surge capacity for severe and critical cases under all of the transmission and testing scenario. There is some capacity available for mild cases, provided documented cases are constrained by testing capacity, albeit, in an event of expanded testing and community transmission, mild cases will also be subjected to deficit of beds. The shortage of beds for even mild and asymptomatic cases can be corroborated with the recent reports from Indian cities of Chennai (21) and Mumbai (22) where government hospitals are running out of beds due to explosion of cases. Many countries are resorting to home isolation of mild and asymptomatic cases, a measure which Union Health Ministry of Indian government has also announced recently. However, we recommend to ramp up institutional capacity to isolate infected cases under institutional care as India’s congested housing conditions are not conducive to quarantining at home. The National Sample Survey Office data (23) reveals that for 60% Indians, per capita space available is less than a single room leading to unprecedented challenges for effective home isolation.

Our model didn’t consider the potential staff shortages in transmission dynamics and capacity readiness due to uncertainty around their predictions and unavailability of quality data. Age-distributed data on severe, critical and recovered cases is rather sparse in India, therefore, we adapted/estimated some parameters from detailed data released by other countries. Modelling is rather a necessary input to guide policy decisions, however, a more comprehensive approach incorporating stakeholder’s analysis, case studies and triangulating information across multitude of sources should be adapted for nuanced decision making. The study can be further extended to map the geographical accessibility of facilities providing COVID-19 care, specifically spatial accessibility for critical care needs to be explored. Also, rapid health facility assessments for COVID-19 preparedness unravelling mean availability of tracer items for emergency response should be conducted for targeted interventions at hospital level. Finally, study can be extended at more granular level to inform demand-supply gap so that resources can be mobilized in commensuration with demand at more local level, thereby enabling local government systems to combat COVID-19 in India.

## Data Availability

Publicly available data is used in the analysis. All the sources and links are mentioned in the study.

## Notes

### Competing Interest Statement

The authors have declared no competing interest.

### Funding Statement

This study is not funded by any agency

